# Global population frequencies of *NAT2* star alleles observed in three large biobanks

**DOI:** 10.64898/2026.06.09.26355281

**Authors:** Katrin Sangkuhl, Michelle Whirl-Carrillo, Mark Woon, Rasika Venkatesh, Karl Keat, Ryan Whaley, Marylyn D. Ritchie, Teri E. Klein

**Author notes:** **Corresponding author:** Teri E. Klein; Department of Biomedical Data Science and Department of Medicine, Stanford University, Stanford, CA, 94303, US.

## Abstract

*NAT2* is an important pharmacogene which encodes the N-acetyltransferase 2 enzyme that is involved in the metabolism of multiple medications, and variants in this gene can affect patient response to these medications. CPIC has published a clinical guideline for prescribing hydralazine using *NAT2* genotypes. Just prior to the guideline, updated *NAT2* star allele numbering and definitions were released, differing somewhat from the historical nomenclature. Clinical pharmacogenomic testing panels often test for the most common star alleles, so knowledge of the most common updated *NAT2* star alleles is critical for the implementation of the CPIC NAT2/hydralazine guideline. We first determine *NAT2* diplotype frequencies from UK Biobank (UKBB) 200k phased genomes, then analyzed allele, diplotype, and phenotype population frequencies from the All of Us Research program, PennMedicine BioBank (PMBB) and UKBB 500k datasets. We found that analyzing NAT2 diplotypes from phased data provides critical information for algorithms designed to predict diplotypes from unphased data. We observed that *NAT2*5*, *\*6*, and *\*4* were the most common star alleles in that order, and the top 11 most frequent *NAT2* star alleles were the same across all biobanks. However, differences in star allele frequencies across biogeographical populations were observed. The largest difference led to a higher frequency of NAT2 poor metabolizer phenotypes as compared to rapid and intermediate metabolizer phenotypes in all global populations except in the EAS population, where NAT2 poor metabolizers were in the minority.

## Introduction

Pharmacogenomics (PGx) is a valuable component of personalized medicine whereby a patient’s genetic test result can help inform the most appropriate medication and dose to maximize efficacy and minimize toxicity. Most clinical PGx panel assays test a limited number of alleles, typically focusing on those believed to be most common in a given population.(1, 2) Since allele frequency is a key factor in determining what is present on a panel, multiple studies have been conducted to determine frequencies in large biobank populations for pharmacogenes included in PGx prescribing guidelines published by experts like the Clinical Pharmacogenetic Implementation Consortium (CPIC).(3–8) CPIC recently published their first guideline to include *NAT2*.(9) This gene also recently transitioned to a slightly different nomenclature system which reclassified many star alleles and contains multiple new allele definitions.(10) The *NAT2* CPIC guideline coupled with the nomenclature transition has created a need to understand the population frequencies of *NAT2* alleles.

The *NAT2* gene encodes N-acetyltransferase 2 (NAT2), a cytosolic phase II xenobiotic-metabolizing enzyme that catalyzes the N-acetylation of arylamine and hydrazine substrates and therefore has a critical role in the detoxification and activation of a wide range of aromatic amines and hydrazine drugs.(11–14) The enzyme is predominantly expressed in the liver(15) and mediates acetylation, a key step in the metabolism of drugs such as isoniazid, procainamide, hydralazine, and sulfonamides.(16–19) NAT2 enzymatic activity is highly variable among individuals due to genetic variation in the *NAT2* gene which results in different acetylator phenotypes.(20–23) These genetic differences have clinical implications, influencing an individual’s susceptibility to drug-induced toxicity and therapeutic efficacy.(18, 24, 25) The study of *NAT2* genetic variation is therefore an important aspect of PGx and CPIC has published a prescribing guideline for hydralazine based on *NAT2* genotypes and phenotypes.(9)

Similar to other pharmacogenes, *NAT2* genotype results are mainly reported using star (*) allele nomenclature which describe haplotypes in the gene. In 2024, *NAT2* nomenclature moved from the original Database of Arylamine N-Acetyltransferases(26, 27) to the Pharmacogene Variation Consortium (PharmVar)(10, 28). PharmVar assigns *NAT2* star alleles by considering the entire coding region of the gene, grouping haplotypes based on unique combinations of amino acids changes and functionally important variants. The PharmVar transition led to a substantial reclassification of *NAT2* star alleles, details of which can be found on the PharmVar website.(10, 28) CPIC assigns a function assignment (e.g. decreased function, increased function) to each *NAT2* star allele, defines NAT2 phenotypes (e.g. poor metabolizer, rapid metabolizer) via allele function combinations and provides actionable dosing recommendations based on phenotypes.(9)

As of January 2026, PharmVar includes definitions for almost 60 *NAT2* star alleles, many of which are various combinations of five common single nucleotide variants (SNVs). These SNVs are rs1801280 (c.341T>C; I114T), rs1799930 (c.590G>A; R197Q), rs1799931 (c.857G>A; G286E), and rs1801279 (c.191G>A; R64Q) which confer decreased NAT2 activity and are found in 40 star alleles in some combination, and rs1208 (c.803G>A; R268K) which is not assumed to alter function and is present in 31 star alleles. The SNVs can be found in numerous combinations *in cis* and *in trans*, leading to multiple possible diplotypes making it difficult to predict the correct diplotype from unphased data. Sometimes, the different diplotype possibilities result in different phenotypes and clinical recommendations (Table 1).

**Table 1.** List of potential ambiguous *NAT2* diplotypes from unphased data and the respective phenotypes. The prioritizated diplotypes based on UKBB 200k phased data are bolded. Ind: Indeterminate; IM: Intermediate Metabolizer; PM: Poor Metabolizer; UKBB: UK Biobank.

| Same phenotype - Diplotype counts available |  |  |  |  |  |  |  |  |
| --- | --- | --- | --- | --- | --- | --- | --- | --- |
| Prioritized Diplotype | Phenotype | Diplotype Count | Alternate Diplotype | Phenotype | Diplotype Count | Alternate Diplotype | Phenotype | Diplotype Count |
| **\*5/\*15** | PM | 2 | **\*6/\*29** | PM | 0 | **\*30/\*46** | PM | 0 |
| **\*7/\*34** | PM | 2 | **\*6/\*40** | PM | 0 |  |  |  |
| **\*5/\*7** | PM | 4,247 | **\*16/\*40** | PM | 0 |  |  |  |
| **\*6/\*14** | PM | 166 | **\*4/\*15** | PM | 0 |  |  |  |
| **\*4/\*5** | IM | 37,270 | **\*1/\*16** | IM | 60 |  |  |  |
| **\*1/\*7** | IM | 80 | **\*4/\*40** | IM | 0 |  |  |  |
| **\*1/\*6** | IM | 749 | **\*4/\*34** | IM | 10 |  |  |  |
| **\*1/\*14** | IM | 63 | **\*4/\*46** | IM | 0 |  |  |  |
| **\*4/\*45** | Ind | 1 | **\*1/\*54** | Ind | 0 |  |  |  |
| **\*6/\*45** | Ind | 3 | **\*34/\*54** | Ind | 0 |  |  |  |
| Different phenotype - Diplotype counts available |  |  |  |  |  |  |  |  |
| Prioritized Diplotype | Phenotype | Diplotype Count | Alternate Diplotype | Phenotype | Diplotype Count | Alternate Diplotype | Phenotype | Diplotype Count |
| **\*5/\*6** | PM | 49,584 | **\*1/\*30** | IM | 1 | **\*16/\*34** | PM | 1 |
| **\*5/\*14** | PM | 211 | **\*4/\*29** | PM | 1 | **\*16/\*46** | IM | 0 |
| **\*6/\*16** | PM | 3,456 | **\*4/\*30** | IM | 1 |  |  |  |
| **\*1/\*36#** | IM | 3 | **\*6/\*44** | Ind | 17 |  |  |  |
| **\*1/\*49#** | IM | 0 | **\*5/\*41** | Ind | 8 |  |  |  |
| **\*6/\*49** | PM | 5 | **\*30/\*41** | Ind | 0 |  |  |  |
| **\*5/\*36** | PM | 17 | **\*30/\*44** | Ind | 0 |  |  |  |
| **\*4/\*49** | IM | 4 | **\*16/\*41** | Ind | 0 |  |  |  |
| **\*4/\*69##** | IM | 1 | **\*5/\*27** | Ind | 0 |  |  |  |
| Same phenotype - No diplotype counts available |  |  |  |  |  |  |  |  |
| Prioritized Diplotype | Phenotype | Allele Counts | Alternate Diplotype | Phenotype | Allele Counts |  |  |  |
| **\*14/\*45** | Ind | 727 - 11 | **\*46/\*54** | Ind | 0 - 0 |  |  |  |
| **\*7/\*45** | Ind | 10,576 - 11 | **\*40/\*54** | Ind | 2 - 0 |  |  |  |
| **\*5/\*54** | Ind | 166,458 - 0 | **\*16/\*45** | Ind | 11,830 - 11 |  |  |  |
| **\*7/\*46** | PM | 10,576 - 0 | **\*14/\*40** | PM | 727 - 2 |  |  |  |
| **\*15/\*16** | PM | 2 - 11,830 | **\*14/\*30** | PM | 727 - 21 |  |  |  |
| Different phenotype - No diplotype counts available |  |  |  |  |  |  |  |  |
| Prioritized Diplotype | Phenotype | Allele Counts | Alternate Diplotype | Phenotype | Allele Counts | Alternate Diplotype | Phenotype | Allele Counts |
| **\*6/\*46** | PM | 119,401 - 0 | **\*14/\*34** | PM | 727 - 66 | **\*1/\*15** | IM | 2,547 - 2 |
| **\*5/\*46** | PM | 166,458 - 0 | **\*1/\*29** | IM | 2,547 - 1 |  |  |  |
| **\*36/\*46** | PM | 43 - 0 | **\*15/\*44** | Ind | 2 - 75 |  |  |  |
| **\*46/\*49** | PM | 0 - 19 | **\*29/\*41** | Ind | 1 - 22 |  |  |  |

Determining the actual combination and diplotype with certainty requires phased data. Most previously published *NAT2* frequency data focus on the five SNVs instead of star alleles, though some recent articles determined star alleles from sequencing data.(3, 4, 29–32) One study used the new PharmVar *NAT2* allele nomenclature but was primarily focused on allele frequencies in people of African descent along with 1000 Genome populations.(3) Our goal was to gain insight into current *NAT2* allele and diplotype distributions across global populations.

We first evaluated UK Biobank (UKBB) 200k phased whole genome sequencing data with the Pharmacogenomics Clinical Annotation Tool (PharmCAT(33, 34)), a PGx bioinformatics tool that analyzes genetic variants to predict diplotypes, phenotypes and drug response to tailor medical treatment to an individual patient’s genetic profile. PharmCAT processes VCF files from next generation sequencing (NGS) or genotyping methods, identifies PGx genotypes and infers star alleles by matching VCF content to established allele definitions sourced from PGx nomenclatures including PharmVar. Given unphased data, the PharmCAT algorithm(35) gives higher scores to predicted alleles with longer matches to the supplied allele defintions which is in line with population data for most genes. For example, *TPMT*1/*3A* is more prevalent than *TPMT*3B/*3C*. However, the longer match does not seem to always be the most prevalent for *NAT2* based on our observations of the UKBB 200k phased data. We therefore adjusted the PharmCAT matching algorithm for unphased *NAT2* variants which result in ambiguous diplotypes using the observed *NAT2* allele frequencies from the UKBB 200k phased data. We then analyzed allele, diplotype, and phenotype population frequencies from unphased data from the All of Us Research program, PennMedicine BioBank (PMBB) and UKBB 500k dataset.

## Materials and Methods

### UKBB 200k phased

The UKBB 200k phased data, located within UKBB field 20279(36), was generated by Ribeiro et al.(37) using a previous release of whole genome sequence data, from UKBB field 24304(38), containing population-level WGS variants for 200k individuals called with GraphTyper. These variants were used as input for the SHAPEIT5 phasing tool(39) in order to generate a phased callset for those 200k individuals. For more details, see UKBB resource 1910(40) in the UKBB showcase.

Computations were done on the UKBB Analysis Workbench (RAP) hosted by DNANexus. We extracted the *NAT2* region on chromosome 8 using bcftools before feeding it to PharmCAT preprocessor using the ‘–missing-to-ref’ flag. All *NAT2* positions used by PharmCAT were covered in this data set. We then used an internally-modified version of PharmCAT that contained the NAT2 allele definition file, referred to as ‘PharmCAT preliminary version 3.0’, to analyze each sample to generate *NAT2* star allele calls with the ‘–reporterCallsOnlyTsv’ flag. The CalcAlleleFrequencies(41) function was used to generate the allele/phenotype frequencies. Resulting diplotypes were evaluated and used to modify PharmCAT’s algorithm for *NAT2* diplotype prioritization for unphased data, released as part of PharmCAT 3.0. The UKBB 200K phased dataset was subsequently rerun on the UK Biobank RAP hosted on DNANexus using the PharmCAT 3.0.1 Docker image (dockerhub: pgkb/pharmcat:3.0.1).

### PharmCAT modification utilizing UKBB 200k phased data

Based on the PharmVar *NAT2* allele definitions, we calculated all possible diplotypes given the same unphased variant input, referred to as “ambiguous calls” (Table1). The list of ambiguous calls was created based on the assumption that all allele positions are in the dataset, as PharmCAT only prioritizes ambiguous calls for samples with 100% coverage. Diplotype counts were assessed in the UKBB 200k phased data to prioritize a single diplotype (first column in Table 1) from each set of ambiguous calls using the following strategy:

- If all ambiguous calls resulted in the same phenotype, select the diplotype with the most observations.
- If ambiguous calls resulted in different phenotypes:

- Select the diplotype with the most observations only if the diplotype is not an Indeterminate phenotype. (Indeterminate phenotypes have no CPIC recommendations and are not actionable.)
- Otherwise, select the diplotype with a non-Indeterminate phenotype. Hence, *NAT2*1/*36* was prioritized over *\*6/*44*, and *NAT2*1/*49* over *\*5/*41*.
- If no ambiguous calls were observed:

- Select the diplotype with a non-Indeterminate phenotype.
- If two diplotypes resulted in the same phenotype, select the diplotype containing the most frequently observed star allele.

Given the prevalence of the five *NAT2* key SNVs, PharmCAT requires input for these positions to determine a diplotype. The modifications for *NAT2* were released in PharmCAT version 3.0 and v3.0.1.(42) Subsequently, PharmCAT v3.0.1 was applied to unphased data from three biobanks.

### All of Us Research Program

Genotypes for 414,830 individuals were acquired using the All of Us v8 release short read whole genome sequencing data. Information on the quality control and other processing of the data can be found in the All of Us Genomic Quality Report(43). We used the pre-prepared exome callset provided in the All of Us Controlled Data Repository. *NAT2* positions were extracted using bcftools. The PharmCAT 3.0.1 docker image (dockerhub: pgkb/pharmcat:3.0.1) was copied into the Google Container Repository for use in All of Us through the dsub job scheduler. The PharmCAT preprocessor was run with the dsub scheduler using the ‘-missing-to-ref’ and ‘-absent-to-ref’ flags. PharmCAT was run in parallel batches on subsets of individuals to generate *NAT2* star allele calls. Resulting calls were analyzed in a Jupyter notebook using python 3.12 to generate allele frequencies. We used the All of Us genetically predicted ancestry auxiliary file to generate allele frequencies in groups defined by their genetic similarity to continental ancestry reference populations in the Human Genome Diversity Project and 1000 Genomes Project(44).

### PennMedicine Biobank (PMBB)

PharmCAT v.3.0.1 was run on Whole Exome Sequencing (WES) data for 57,170 individuals from the PMBB 3.0 release(45), consisting of 43,913 samples from the previous Freeze 2.0 and 13,257 new samples sequenced in 2024. Genetically inferred ancestry (GIA) was calculated by Regeneron Genetics Center (RGC) by projecting the study participants onto HapMap3 principal components (PCs) from the Broad Institute and modeling PC distributions per ancestral group using 2D kernel density estimation (KDE) (Figure S1). Variant calling and readapting across both freezes were also performed by RGC. *NAT2* positions were extracted from VCFs using bcftools. We ran the PharmCAT preprocessor using the ‘-missing-to-ref’ and ‘-absent-to-ref’ flags. PharmCAT was run in parallel batches on subsets of individuals to generate *NAT2* star allele calls and report files. Allele frequency per ancestry group was calculated by generating a calls report tsv file using the ‘-reporterCallsOnlyTsv’ flag and then running the CalcAlleleFrequencies(41) function on this file.

### UK Biobank (UKBB) 500k unphased

We used the 2023 release of individual-level WGS data for 500k individuals, called using DRAGEN(46). This data is available at UKBB field 24310(47) and more information on data processing, quality control and other procedures used to create this dataset are described in a recent publication(48). Custom scripts were used to extract known PGx regions from these VCF files, which were used as input for the PharmCAT preprocessor and PharmCAT. All computations were done on the UK Biobank RAP hosted on DNANexus using the PharmCAT 3.0.1 Docker image (dockerhub: pgkb/pharmcat:3.0.1). We extracted the *NAT2* region on chromosome 8 using bcftools before feeding it to the PharmCAT preprocessor using the ‘–missing-to-ref flag’. PharmCAT was used to analyze each sample to generate *NAT2* star allele calls with the ‘–reporterCallsOnlyTsv’ flag. Finally, CalcAlleleFrequencies(41) was used to generate the allele/phenotype frequencies.

We inferred genetic ancestry in the UKBB using the genotype data (UKB field 22418) to perform joint principal component analysis (PCA) with the 1000 Genomes Phase 3 release data (1KG) (Figure S2). We assigned the UKBB participants to superpopulations based on their genetic similarity to the superpopulation groupings in the 1KG reference. We used PLINK 1.9.0-b.8 to apply a 0.05 minor allele frequency cutoff, applied a HWE p-value threshold of 1e-6, subset to autosomal variants, and performed LD pruning using ‘--indep-pairwise 50 5 0.5’ on the genotype data. The set of shared variants between the genotype data and the 1KG data were then jointly run on smartpca (version 18140, eigensoft 8.0.0) in fastmode. The 1KG PCA eigenvectors were then used to train a quadratic discriminant analysis classifier using the R MASS package (version 7.3-60), which was then applied to the UKBB genotype eigenvectors to infer ancestry.

### Allele function and phenotype mapping

To derive NAT2 phenotype frequencies, we applied CPIC phenotype definitions and allele function assignments(9) to (1) PharmCAT diplotype output for each biobank and (2) *NAT2* diplotypes provided by Malinga *et al*.(3) in the Supplementary Table 5. A Poor Metabolizer (PM) has two decreased function alleles. An Intermediate Metabolizer (IM) has one decreased function allele and one increased function allele. A Rapid Metabolizer (RM) has two increased function alleles. Any diplotype that includes an uncertain function or unknown function allele is an Indeterminate phenotype.

## Results

### UKBB 200k phased data

The 48 *NAT2* alleles in PharmCAT v3.0.1 are defined by 37 variant positions. All positions were covered in this data set. Table 1 lists the 27 ambiguous *NAT2* diplotype calls that are possible with unphased *NAT2* variant positions and the resulting counts for diplotypes found in the UKBB 200k phased data set. PharmCAT’s Named Allele Matcher algorithm in v3.0.1 was modified for *NAT2* as described in the methods section based on this data. All allele, diplotype, and phenotype frequencies for the UKBB 200k phased data are available in Table S1.

### Biobank population groups

Biobank datasets include individuals of African (AFR), Admixed American (AMR), South Asian (SAS), East Asian (EAS), and European (EUR) descent. The All of Us data also include individuals of Middle Eastern (MID) descent. The remaining samples for each biobank were combined in the ‘Other’ population group. Complete frequency data for each population in each biobank is available in the supplements (Table S1). Table 2 summarizes the number of alleles covered and identified per population group and biobank. The EUR population had the largest sample number in all biobanks, however the All of Us data also have larger AFR and AMR cohorts than the other biobanks. The EAS and SAS populations have the least number of alleles identified. AFR and EUR populations display more diverse allele representation.

**Table 2.**
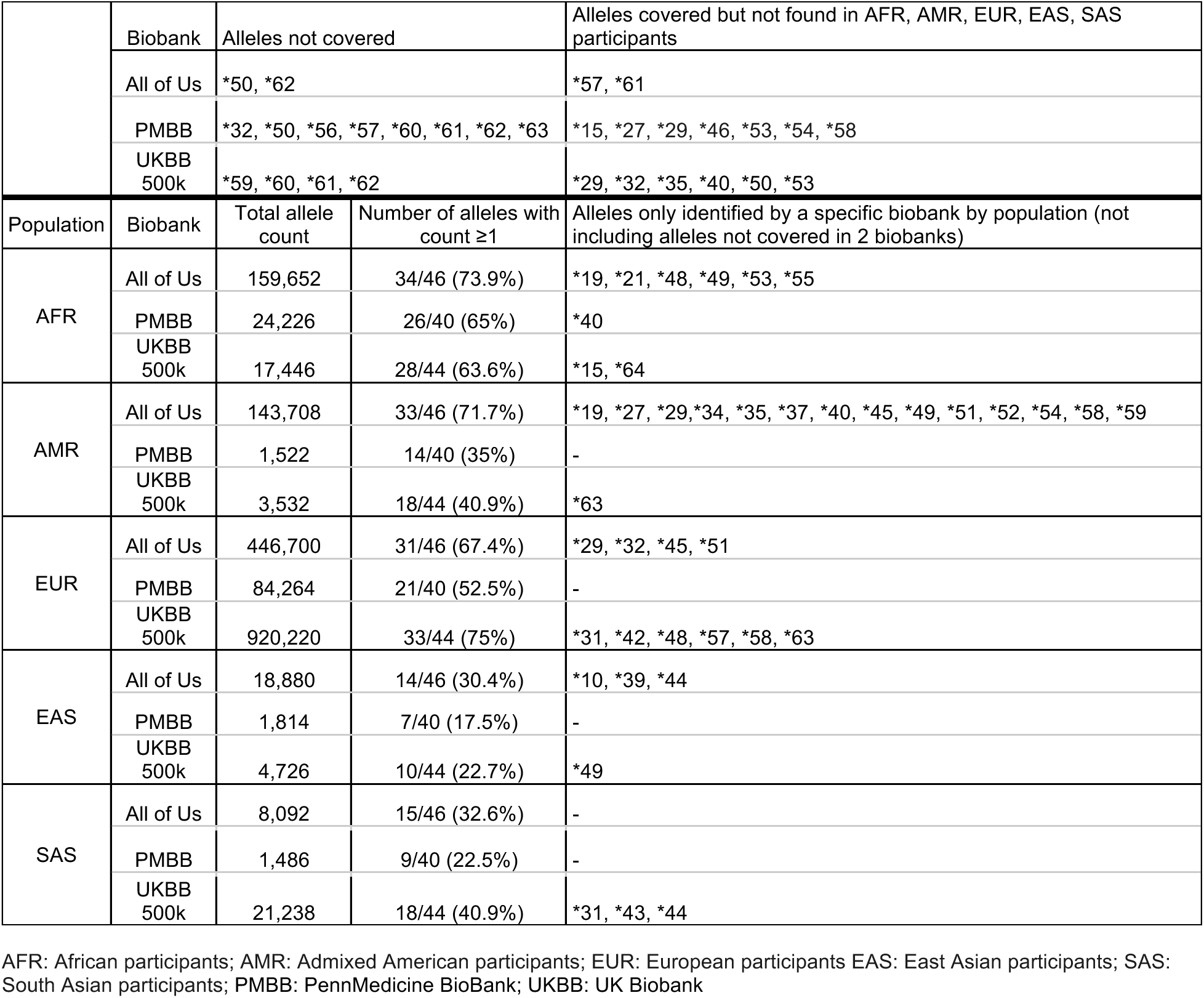
Summary of the *NAT2* alleles identified by population.

### Unphased biobank data

All allele, diplotype, and phenotype frequency results by population group for the All of Us, PMBB, UKBB 500k unphased data are found in Tables S1.

Of the 37 *NAT2* variant positions in PharmCAT, two positions were missing from the All of Us data: chr8:18400010 (found in *\*50*) and chr8:18400539 (found in \**62*), leaving 46 detectable star alleles. Eight positions/alleles were missing in the PMBB data: chr8: 18400010 (*\*50*), chr8:18400061 (*\*56*), chr8:18400206 *(*32*), chr8:18400324 (*\*57*), chr8:18400494 (*\*60*), chr8:18400509 (*\*61*), chr8:18400539 (*\*62*), and chr8:18400653 (*\*63*), leaving 40 detectable star alleles. Four positions/alleles were missing in the UKBB 500k unphased data: chr8:18400478 (*\*59*), chr8:18400494 (*\*60*), chr8:18400509 (*\*61*) and chr8:18400539 (*\*62*), leaving 44 detectable star alleles. Of the detectable star alleles in each biobank, some were not observed in any sample: *\*57* and *\*61* (only found in MID, Table S1) in All of Us; *\*15*, *\*27*, *\*29*, *\*46*, *\*53*, *\*54,* and *\*58* in PMBB; and *\*29*, *\*32*, *\*35*, *\*40*, *\*50,* and *\*53* in the UKBB 500k unphased data. Table 2 provides an overview of alleles not covered in each biobank dataset, as well as which alleles were detectable but not observed.

Based on overall population frequency, the 11 most frequent alleles were the same across all three unphased datasets, with *NAT2*5*, *\*6*, and *\*4* the top three most common alleles in that order (Table S2). The exact order of the next four most frequent alleles differed slightly across biobanks. For example, the frequency of *NAT2*14* is slightly higher in the All of Us data compared to the other biobanks, largely because it is more prevalent in the AFR population which is better represented in that dataset.

For the remaining alleles that were found in all three biobanks, the frequencies by allele were comparable within each population (Figure 1, Table S3). PharmCAT requires coverage for the positions found in *NAT2*4*, *\*5*, *\*6*, *\*7*, and *\*14*. Those alleles were identified in all biobanks and populations with the exception of *\*14*, which was not present in the EAS and SAS populations in the PMBB. In the All of Us and UKBB 500k unphased data, *NAT2*14* showed a lower frequency in EAS and SAS poulations as compared to AFR, AMR, EUR populations. More details about individual star allele frequencies, including very rare alleles, can be found in the Supplementary Materials.

**Figure 1.**
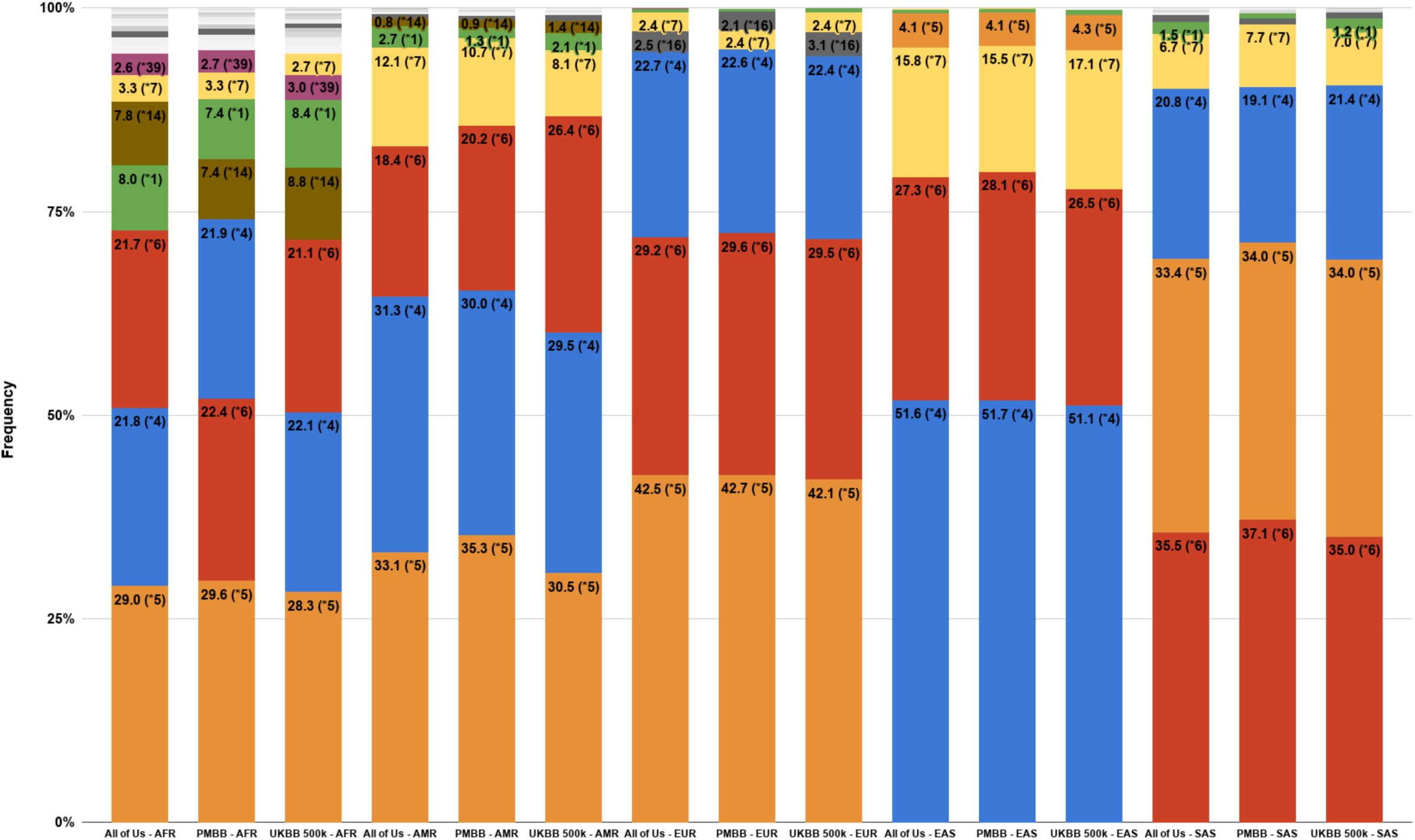
*NAT2* allele frequencies. Highest allele frequencies by population group in All of Us, PMBB, and UKBB 500k unphased data. AFR: African participants; AMR: Admixed American participants; EUR: European participants EAS: East Asian participants; SAS: South Asian participants; PMBB: PennMedicine BioBank; UKBB: UK Biobank.

Three hundred thirty-four samples in the All of Us data, one sample in the PMBB data and one sample in the UKBB 500k unphased data had ambiguous calls due to missing variant information at one or more *NAT2* variant positions. Some samples could not be matched to any known *NAT2* diplotype (labeled “unknown”) because the sample: 1) contained an unrecognized nucleotide change (e.g. PharmVar documented a G>T change at a position, and the sample contained a G>A change), 2) was missing at least one of the five PharmCAT-required positions, or 3) contained a combination of variants not found in the PharmVar *NAT2* allele definitions (see Table S4).

### Predicted phenotype frequencies

Predicted phenotype frequencies are consistent across biobank population groups (Table 3). For all populations except EAS, the PM phenotype is most frequent, followed by IM then RM. The difference between PM and IM frequencies is more pronounced in the EUR and SAS populations as compared to the AFR and AMR populations. In the EAS population, the IM phenotype is most frequent, followed by RM then PM. The phenotype frequencies align with the allele function frequencies, since the increased function allele *\*4* is the most prevalent allele in the EAS population (Figure 2). For the AFR and AMR populations, *NAT2*4* is the second most frequent allele, while for the EUR and SAS populations, it is third after *\*5* and *\*6*.

**Figure 2.**
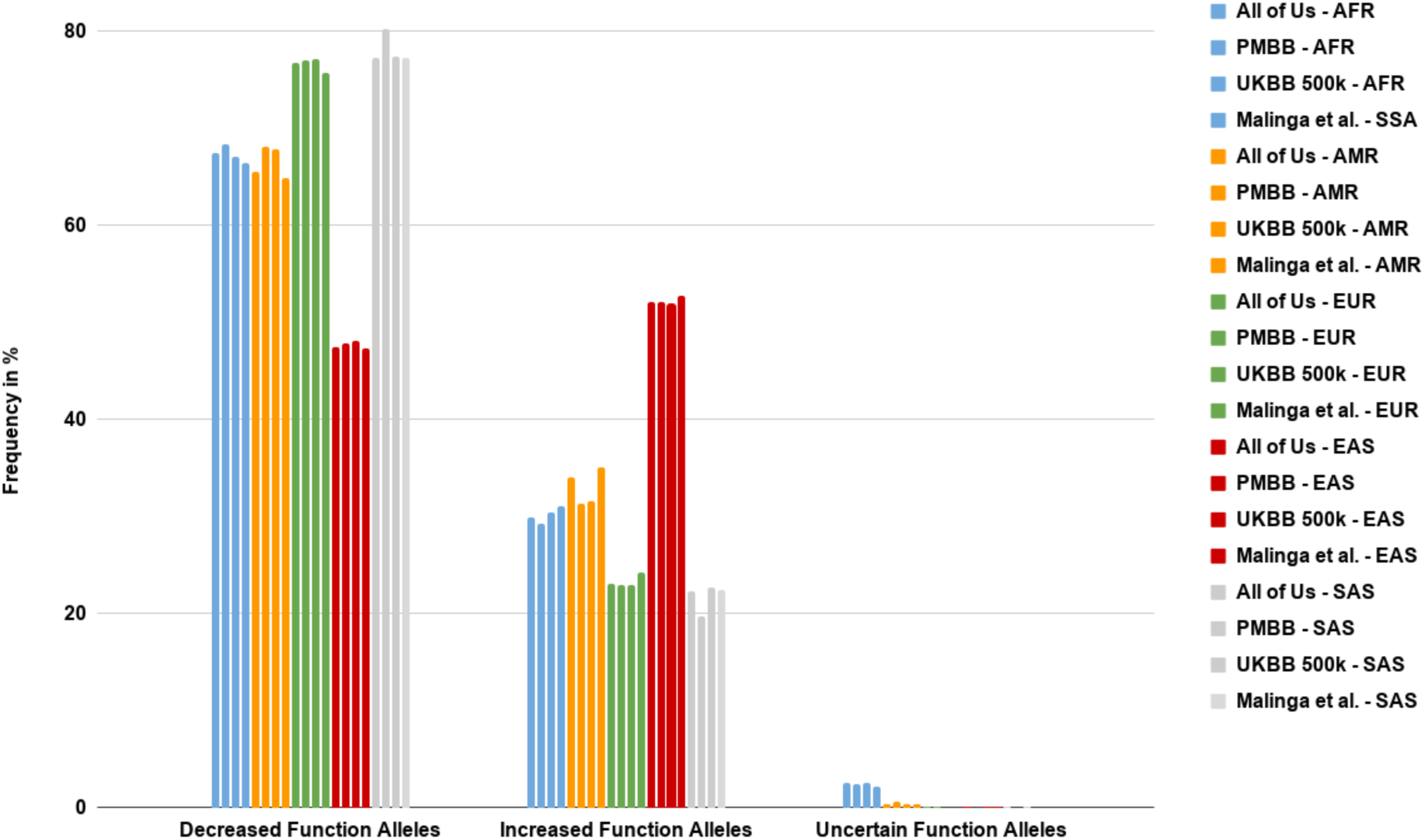
*NAT2* allele frequencies grouped by function. Frequencies are shown by population group as found in All of Us, PMBB, UKBB 500k unphased data and what was reported in Malinga *et al*., 2025(3). AFR: African participants; AMR: Admixed American participants; EUR: European participants EAS: East Asian participants; SAS: South Asian participants; PMBB: PennMedicine BioBank; UKBB: UK Biobank.

**Table 3.** NAT2 phenotype frequency by population and biobank. NAT2 phenotype frequencies are based on CPIC phenotype definitions and allele function assignments (9) and applied to PharmCAT diplotype output for each biobank and *NAT2* diplotypes provided by Malinga *et al*., 2025(3). AFR: African participants; AMR: Admixed American participants; EUR: European participants EAS: East Asian participants; SAS: South Asian participants; PMBB: PennMedicine BioBank; UKBB: UK Biobank

|  | AFR Frequency % |  |  | Malinga *et al.* 2025 |  |
| --- | --- | --- | --- | --- | --- |
| Phenotype | All of Us<br>(n=79,826) | PMBB<br>(n=12,113) | UKBB 500k<br>(n=8,727) | African American/<br>Afro Caribbean<br>(n=157) | Sub-Saharan<br>African<br>(n=1079) |
| NAT2 Indeterminate | 4.5 | 4.1 | 4.9 | 3 | 4.2 |
| NAT2 Rapid Metabolizer | 8.9 | 8.2 | 8.9 | 8.3 | 8.5 |
| NAT2 Intermediate Metabolizer | 40.9 | 41.4 | 41.6 | 42.3 | 43.5 |
| Rapid + Intermediate | <u>49.8</u> | <u>49.6</u> | <u>50.5</u> | <u>50.6</u> | <u>52</u> |
| NAT2 Poor Metabolizer | <u>45.5</u> | <u>46.3</u> | <u>44.5</u> | <u>45.6</u> | <u>43.3</u> |
|  | AMR Frequency % |  |  |  |  |
| Phenotype | All of Us<br>(n=71,854) | PMBB<br>(n=761) | UKBB 500k<br>(n=1,766) | Malinga *et al.* 2025 (n=347) |  |
| NAT2 Indeterminate | 0.5 | 0.8 | 0.7 | 0.6 |  |
| NAT2 Rapid Metabolizer | 11.9 | 9.6 | 10.8 | 14.1 |  |
| NAT2 Intermediate Metabolizer | 44 | 43.5 | 41.5 | 41.6 |  |
| Rapid + Intermediate | <u>55.9</u> | <u>53.1</u> | <u>52.3</u> | <u>55.7</u> |  |
| NAT2 Poor Metabolizer | <u>43.3</u> | <u>46.1</u> | <u>46.8</u> | <u>43.9</u> |  |
|  | EUR Frequency % |  |  |  |  |
| Phenotype | All of Us<br>(n=223,350) | PMBB<br>(n=42,133) | UKBB 500k<br>(n=460,113) | Malinga *et al.* 2025 (n=503) |  |
| NAT2 Indeterminate | 0.1 | 0.1 | 0 | 0 |  |
| NAT2 Rapid Metabolizer | 5.4 | 5.2 | 5.3 | 6.8 |  |
| NAT2 Intermediate Metabolizer | 35.5 | 35.5 | 35.2 | 34.8 |  |
| Rapid + Intermediate | <u>40.9</u> | <u>40.7</u> | <u>40.5</u> | <u>41.6</u> |  |
| NAT2 Poor Metabolizer | <u>58.9</u> | <u>59.2</u> | <u>59.4</u> | <u>58.2</u> |  |
|  | EAS Frequency % |  |  |  |  |
| Phenotype | All of Us<br>(n=9,440) | PMBB<br>(n=907) | UKBB 500k<br>(n=2,364) | Malinga *et al.* 2025 (n=504) |  |
| NAT2 Indeterminate | 0.1 | 0 | 0 | 0.2 |  |
| NAT2 Rapid Metabolizer | 28.2 | 29 | 26.9 | 30.9 |  |
| NAT2 Intermediate Metabolizer | 47.7 | 46.2 | 49.6 | 43.4 |  |
| Rapid + Intermediate | <u>75.9</u> | <u>75.2</u> | <u>76.5</u> | <u>74.3</u> |  |
| NAT2 Poor Metabolizer | <u>23.6</u> | <u>24.7</u> | <u>23.3</u> | <u>25.6</u> |  |
|  | SAS Frequency % |  |  |  |  |
| Phenotype | All of Us<br>(n=4,046) | PMBB<br>(n=743) | UKBB 500k<br>(n=10,632) | Malinga *et al.* 2025 (n=489) |  |
| NAT2 Indeterminate | 0.1 | 0 | 0 | 0 |  |
| NAT2 Rapid Metabolizer | 5.5 | 4.2 | 5.4 | 5.9 |  |
| NAT2 Intermediate Metabolizer | 33.5 | 31.1 | 34.2 | 33.1 |  |
| Rapid + Intermediate | <u>39.1</u> | <u>35.3</u> | <u>39.7</u> | <u>39</u> |  |
| NAT2 Poor Metabolizer | <u>60.5</u> | <u>64.6</u> | <u>60.1</u> | <u>60.7</u> |  |

The CPIC NAT2-hydralazine guideline gives the same recommendations for RM and IM phenotypes, and different guidance for the PM phenotype(9), essentially combining the RM and IM phenotypes into one group. For the AFR and AMR populations, grouping RM and IM together results in a slightly higher frequency than the PM phenotype. However, for the EUR and SAS populations, the PM phenotype is more frequent than RM and IM together. The EAS population has the lowest frequency of PM phenotypes of all the population groups at around 25%. Overall, the biobank phenotype frequencies show high concordance with phenotype frequencies derived from published data by Malinga *et al*.(3) which was comprised of diplotypes called from 1000 Genomes and a Sub-Saharan African population using alternate software (Table 3).

## Discussion

The numerous combinations of the five common variants found in the *NAT2* star allele definitions (rs1801280 (c.341T>C; I114T), rs1799930 (c.590G>A; R197Q), rs1799931 (c.857G>A; G286E), and rs1801279 (c.191G>A; R64Q)) which confer decreased NAT2 activity and are found in 40 star alleles in some combination, together with rs1208 (c.803G>A; R268K)), cause substantial difficulty for *NAT2* diplotype prediction from unphased data. Out of the 48 star alleles included in PharmCAT v3.0.1, 40 star alleles (83%) include at least one of the five SNVs. Of those 40 star alleles, 20 include two of the variants, and two include three of the variants (Table S5). The amount of overlap between key SNVs and star alleles presented a new challenge for the PharmCAT algorithm, which defaulted to the diplotype that matches the most variant positions on one allele (*in cis*). Before the addition of *NAT2*, the “longest match” approach had been largely successful for pharmacogenes. For example, *TMPT*3A* is comprised of two variants, one of which is found in *TPMT*3B* and the other in *TMPT*3C*. In unphased data when both variants are heterozygous, the variants could be *in trans* (*TPMT*3B/TPMT*3C*) or *in cis* (*TPMT*1/TPMT*3A*). *TMPT*3A* is more common than *TPMT*3B* or *TMPT*3C* and *TPMT*1/*3A* is more common than *TPMT*3B/TPMT*3C* (see ClinPGx TPMT resources(49)) so PharmCAT’s algorithm of matching the most variants *in cis* was appropriate and successful.

However, historical frequency data suggested that this approach may not work for *NAT2*.(10) We applied PharmCAT to UKBB 200k phased whole genome sequencing data to determine *NAT2* allele and diplotype frequencies which verified that the longest match approach was not appropriate in some situations. We then altered PharmCAT’s approach to unphased *NAT2* to favor the allele/diplotype with the highest frequency when multiple calls are possible. We made exceptions to this rule in cases where neither of the diplotypes was common and one diplotype resulted in a non-actionable phenotype according to CPIC. In these cases, we defaulted to the actionable diplotype (Table 1).

Papanikolaou *et al*. recently published a review with a detailed discussion of *NAT2* genetic variation frequencies across world populations that were inferred from the small set of common SNVs.(10) In general, the allele frequency results from the biobank analyses presented here are consistent with this review. However, some differences were apparent. For example, while the biobank frequencies for *5 in the EUR and AFR populations were largely in the 30-50% range given in the review, it was consistently over 40% in the EUR population while ≤30% in the AFR population. The *NAT2*1* frequency in the AMR and SAS populations in the biobank data (generally >1%) are markedly different from the EUR and EAS populations (<0.7%). The *\*6* frequency is ∼30% or higher in the EUR and SAS populations rather than the ∼20% given in the review. The largest difference between the review and biobank datasets is that the review groups the EAS and SAS populations together, while our biobank data shows a pronounced difference between those two populations (Table S6) across star alleles. For *\*4*, the frequency in EAS population is >50% but in the biobanks the frequency in the SAS population is less than half of that at ∼20%, much more similar to the AFR, AMR and EUR populations. Again for *\*7*, the frequency for the EAS population is 16-17%, but for the SAS population, the frequency is cut by half to 7-8%. Additionally, the historical data based on the fie SNVs cannot account for the many alleles that do not contain these variants, including alleles recently deposited in PharmVar.

Several articles have been published on *NAT2* frequencies using sequencing data.(4, 29–32) Malinga *et al*. used the new PharmVar *NAT2* allele nomenclature focusing on allele frequencies in people of African descent along with 1000 Genome populations.(3) Comparing diplotypes from the three biobanks to those presented by Malinga *et al*. shows a difference in assigning c.341T>C and c.803G>A together (*\*16*) vs on two different strands (*\*4/*5*) (Table S7). Malinga *et al*. determined a much higher frequency of *\*1/*16* vs *\*4/*5*, *\*16/*34* vs *\*5/*6*, and *\*16/*40* vs *\*5/*7*, while our biobank results based on phased data frequencies are reversed. In fact, *\*16/*40* was not observed at all in the UKBB 200k phased dataset, *\*16/*34* was seen only one subject and *\*1/*16* was seen in only 120 subjects as compared to <74,000 subjects with *\*4/*5*. However, after translating the diplotypes from Malinga *et al*. into phenotypes, their data align with our phenotype frequencies (Figure 2 and Table 3). Even though the phenotype frequencies are similar, the divergent diplotypes result in significant star allele frequency differences. The *Malinga et al.* dataset puts the *\*16* allele frequency at 10.8%, much higher than the 0.7% seen in the AFR population in the phased biobank data, and greatly decreases the *\*5* and *\*4* frequencies (17.5% and 15.2%) found in the AFR population in the phased data (41.2% and 22.6%).

A limitation of the diplotype prioritization approach based on the UKBB 200k phased data is the comparatively small sample numbers in non-EUR populations. Furthermore, several new star alleles have been deposited into PharmVar after the PharmCAT runs. Next steps include using an updated version of PharmCAT with the addition of the most recent *NAT2* star alleles (PharmCAT v3.1.1) on the newly available UKBB 500k phased dataset to help fine tune diplotype prioritization. In addition, future long read sequencing results could be very valuable since they do not rely on a computational phasing methods which could be biased or inaccurate for some populations.

The results presented here highlight the importance of large and diverse population datasets to cover a more comprehensive number of star alleles and the impact of phasing on *NAT2* alleles. Our findings (1) demonstrate the ambiguity of *NAT2* diplotype calling using unphased data, (2) largely corroborate historical frequency data for the most common *NAT2* star alleles but reveal important detailed population differences, and (3) determine global population frequencies for newly identified and reclassified PharmVar *NAT2* star alleles. These data can be found on the ClinPGx website(50) and can be potentially used by laboratories and PGx guideline groups to help prioritize which star alleles and variants to add to PGx testing panels in the future.

## Supplemental Information

The Supplementary Material includes Further Results section, two figures, and seven tables.

## Data Availability

All data produced in the present work are contained in the manuscript, accessible in the supplements or at https://www.clinpgx.org/downloads.

## Acknowledgments

This work was funded by the National Institutes of Health (NIH) for PharmCAT/CPIC (U24HG013077) and PharmGKB (U24HG010615). We gratefully acknowledge All of Us participants for their contributions, without whom this research would not have been possible. We also thank the NIH’s All of Us Research Program for making available the participant data examined in this study. We acknowledge the PennMedicine BioBank (PMBB) for providing data and thank the patient-participants of Penn Medicine who consented to participate in this research program. We would also like to thank the PennMedicine BioBank team and Regeneron Genetics Center for providing genetic variant data for analysis. The PMBB is approved under IRB protocol# 813913 and supported by Perelman School of Medicine at University of Pennsylvania, a gift from the Smilow family, and the National Center for Advancing Translational Sciences of the National Institutes of Health under CTSA award number UL1TR001878. We acknowledge all the participants of the UK Biobank. The use of the UK Biobank resources was approved under Application ID 703474.

## Conflicts of interests

M.D.R. is a Bioinformatics Consultant for AbbVie. All other authors declare no competing interests.

## Author contributions

K.S., M.W.-C., M.W., K.K., R.V, T.E.K., and M.D.R. wrote the manuscript. K.S. and M.W.-C. designed the research. M.W., K.K., R.V. and R.W. contributed analytical tools. K.S., M.W.-C., M.W., K.K. and R.V performed the research and analyzed the data.

## Web resources

ClinPGx, https://www.clinpgx.org

PharmCAT, https://pharmcat.clinpgx.org

PharmCAT GitHub, https://github.com/PharmGKB/PharmCAT

PharmCAT DockerHub, https://hub.docker.com/r/pgkb/pharmcat

PharmVar, https://www.pharmvar.org

## Data and code availability

The allele, diplotype and phenotype frequency datasets are available in the supplementary material and online on the ClinPGx website (http://www.clinpgx.org/downloads). The PharmCAT code is available online through the PharmCAT website (http://pharmcat.clinpgx.org).

## Notes

**Funding:** This work was funded by the National Institutes of Health (NIH) for PharmCAT/CPIC (U24HG013077) and PharmGKB (U24HG010615).

### Author Declarations

The use of the UK Biobank resources was approved under Application ID 703474. Data used for determing frequencies in this study are available on the All of Us Researcher Workbench. The PennMedicine Biobank is approved under IRB protocol# 813913.

