## Supplemental Material for "Global population frequencies of *NAT2* star alleles observed in three large biobanks"

### Further Results

*NAT2*1* – assigned when all covered positions are reference - was also identified in all biobanks and all populations. *NAT2*16* was present in all except the UKBB 500k unphased data EAS population. Among all the identified alleles, *NAT2*5* is the most prevalent in AFR, AMR, EUR populations, second most frequent in the SAS population after *6 and fourth most frequent in the EAS population after **4*, **6*, and **7*. In the AFR population in all biobanks, **4* and **6* are around 22%, while in the AMR population, **4* (29-30%) is more frequent than **6* (close to 20%) except for the UKBB 500k unphased data which had less divergent frequencies for both alleles. In the EUR population, the order is reversed with **6* around 30% and **4* around 22%. In the SAS population, **4* has also a frequency of around 20%, while **6* has the highest frequency at 35.5-37% - the highest among all populations. The EAS population displays a unique allele profile compared to the other populations with *4 making up 50% of the alleles, **6* frequency 26-28%, followed by **7* with a frequency of 15-16% - the highest among all populations (AFR around 3%, AMR 8-12%, EUR 2-2.5%, and SAS around 7%). Among the most tested *NAT2* alleles with decreased function *(*5*, **6*, **7*, **14*), **14* is the least frequent, found in AFR around 7-9%, AMR 0.8-1.4%, EUR 0.02-0.04%, EAS 0.01-0.02%, and SAS 0.03-0.04%. *NAT2*1* is less frequent than **4*, and found at 7-

9% in AFR, 1-3% in AMR, 0.3-0.5% in EUR, 0.4-0.7% in EAS, and 0.7-1.5% in SAS. *NAT2*16* is assigned decreased function since it is contains 341T>C (the defining variant of **5*) and also includes 803G>A (the defining variant of **4*). In unphased data, the frequency of **16* is around 0.7% in AFR, 0.5-0.6% in AMR, around 2.5% in EUR, around 4% in EAS, and 0.6-0.75% in SAS. This was not much different from the **16* frequency observed in the UKBB 200k phased data samples: 0.7% in AFR, 0.6% in AMR, around 3% in EUR, and 0.8% in SAS. However, *16 was not observed at all in the UKBB 200k phased data EAS population.

*NAT2*61* was not covered in PMBB and UKBB 500k unphased data and not found in any of the included populations from the All of Us data.

*NAT2*57* was not covered in PMBB, not found in any population in the All of Us data and only in the UKBB 500k unphased data EUR population.

The alleles **46* and **53* were covered in all 3 biobanks but only identified in the AFR populations (**46*: allele count 5 in All of Us and 2 in UKBB; **53*: allele count 12 in All of Us).

**Figures**

Figure S1. Genetically inferred ancestry (GIA) across the PennMedicine BioBank (PMBB) cohort. Principal component analysis (PCA) of the PMBB cohort, with GIA assigned by projection onto the HapMap3 reference panel. Individuals are colored by inferred ancestry.

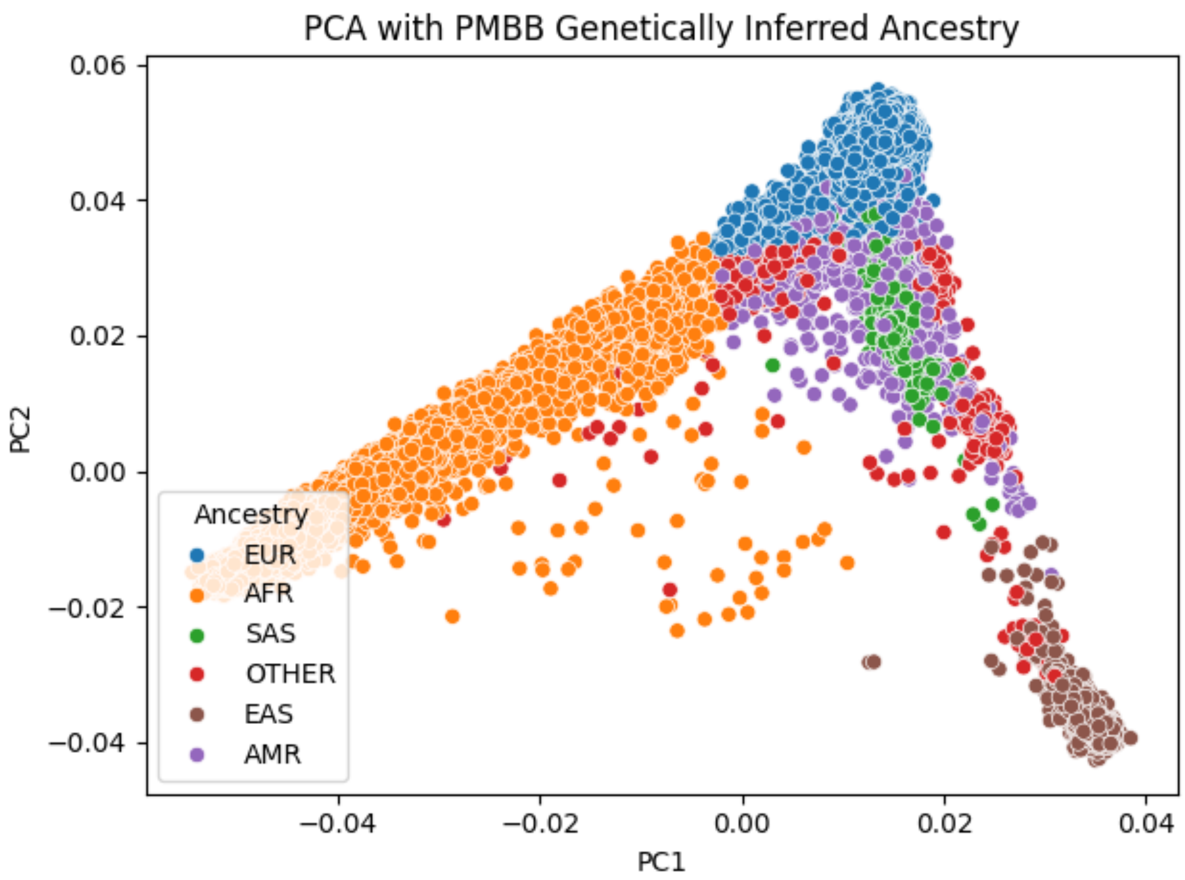

Figure S2. Genetically inferred ancestry (GIA) across the UK Biobank (UKBB) cohort. Principal component analysis (PCA) of the UKBB cohort, with GIA assigned by projection onto the 1000 Genomes (1KG) reference panel. Individuals are colored by inferred ancestry.

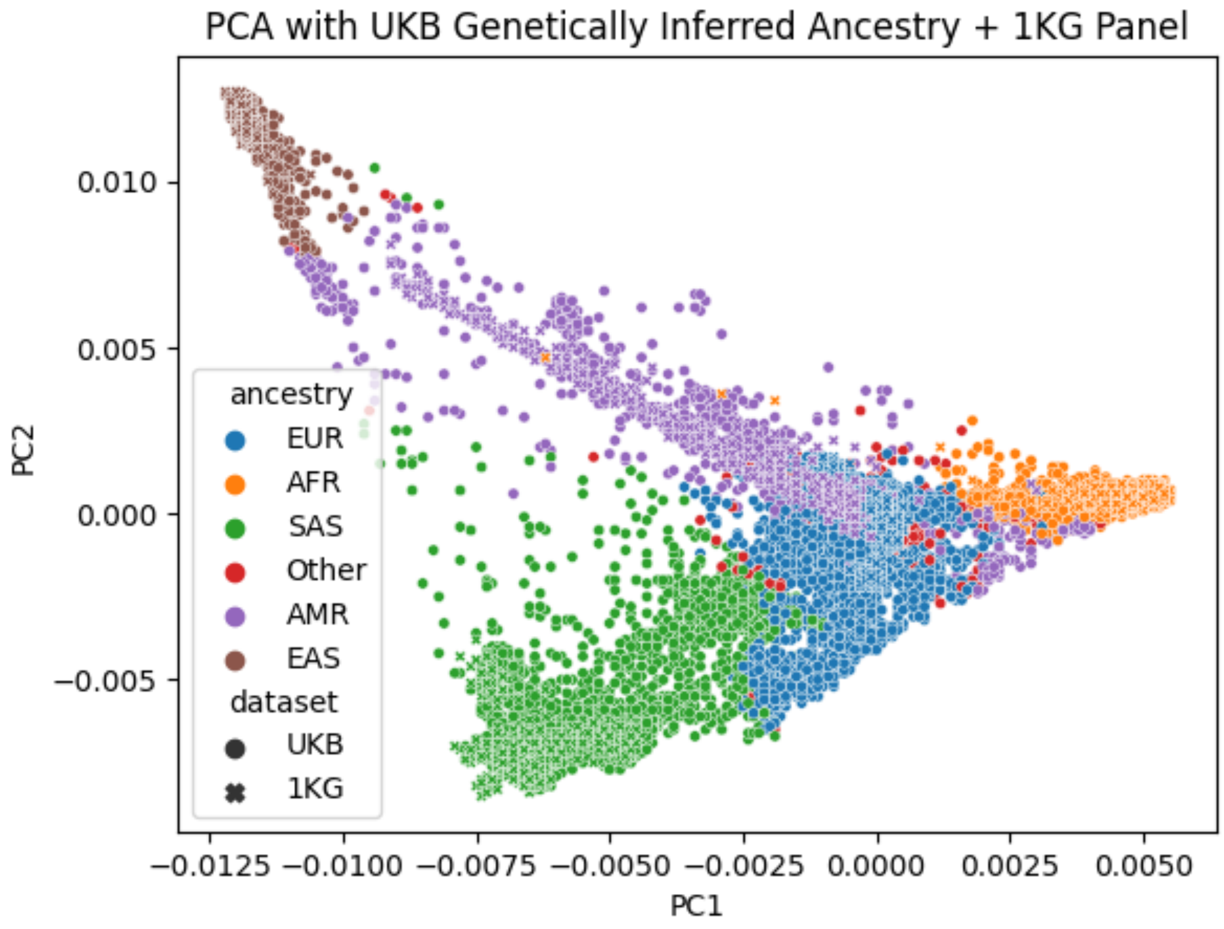

**Tables**

Tables S1. NAT2 allele, diplotype, and phenotype frequencies in the UK Biobank 200k phased, All of Us Research program, PennMedicine BioBank, and UK Biobank 500k unphased dataset. Frequencies are analyzed in 2025 with PharmCAT v3.0.1. Alleles with frequency of zero (position covered but allele not found) are not included.

Abb.: AFR – African participants; AMR – admixed American participants; EUR – European participants; EAS – East Asian participants; SAS – South Asian participants; PMBB – PennMedicine Biobank; UKBB – UK Biobank

Available on the ClinPGx downloads page, section: Other Datasets - NAT2 Biobank Frequencies

Table S2. Overall *NAT2* allele frequencies identified in All of Us, PennMedicine BioBank, UK Biobank 500k unphased datasets (analyzed with PharmCAT v3.0.1). The top 11 most frequent alleles across biobanks are highlighted in yellow. Alleles with frequency of zero (position covered but allele not found) are not included.

Abb.: AFR – African participants; AMR – admixed American participants; EUR – European participants; EAS – East Asian participants; SAS – South Asian participants; PMBB – PennMedicine Biobank; UKBB – UK Biobank

Available on the ClinPGx downloads page, section: Other Datasets - NAT2 Biobank Frequencies

Table S3. *NAT2* allele frequencies by population in the All of Us, PennMedicine BioBank, and UK BioBank 500k unphased datasets (analyzed with PharmCAT v3.0.1). Alleles with frequency of zero (position covered but allele not found) are not included.

Abb.: AFR – African participants; AMR – admixed American participants; EUR – European participants; EAS – East Asian participants; SAS – South Asian participants; PMBB – PennMedicine Biobank; UKBB – UK Biobank

Available on the ClinPGx downloads page, section: Other Datasets - NAT2 Biobank Frequencies

Table S4. Summary of the unknown call counts per biobank.

Abb.: PMBB – PennMedicine Biobank; UKBB – UK Biobank; rsID – rs Identifier

| Biobank | All of Us | PMBB | UKBB 500k |
|---|---|---|---|
| No. of samples without a call | **518** | **28** | **116** |
| No. of samples missing a required position | **278** | **5** | **33** |
| Missing required positions (number) | rs1208 (22)<br>rs1801279 (45)<br>rs1801280 (127)<br>rs1799930 (51)<br>rs1799931 (31)<br>rs1208, rs1799931 (1)<br>rs1208, rs1799930 (1) | rs1208 (3)<br>rs1801279 (2) | rs1208 (13)<br>rs1801280 (14)<br>rs1799930 (5)<br>rs1799931 (1) |
| No. of samples with undocumented position | **94** | **5** | **37** |
| Undocumented positions (number) | rs561124342 (2)<br>rs375746304 (4)<br>rs1801280 (1)<br>rs72466456 (64)<br>rs4986996 (6)<br>rs72554617 (4)<br>rs1801279 (1)<br>rs79050330 (3)<br>rs72466458 (1)<br>rs561124342 (5)<br>rs549917500 (2)<br>rs149283608 (1) | rs561124342 (2)<br>rs375746304 (1)<br>rs1801280 (1)<br>rs45477599 (1) | rs561124342 (1)<br>rs375746304 (1)<br>rs72466456 (10)<br>rs4986996 (4)<br>rs72554617 (8)<br>rs1801279 (2)<br>rs79050330 (1)<br>rs72466458 (1)<br>rs12720065 (2)<br>rs139351995 (1)<br>rs532310930 (1)<br>rs72466457(1)<br>rs45618543 (4) |
| No. of samples with novel combinations | **146** | **18** | **46** |
| Examples of novel combinations of NAT2 PGx positions<br>(For samples where several diplotype options are possible due to unphased data, one diplotype per variant combination is chosen, e.g., *19/[*5+*6] OR [*5+*19]/*6)<br>^[*5+g.18400592C>T] = *69 | *16/[g.18400841G>A]<br>*16/[g.18400349G>A]<br>*5/[*5+g.18400592C>T^]<br>*6/[*5+g.18400592C>T^]<br>*5/[*5+*6]<br>*19/[*5+*6]<br>*4/[*4+g.18400841G>A]<br>*4/[*4+g.18400349G>A]<br>*4/[g.18400841G>A]<br>*5/[*5+g.18400841G>A]<br>*6/[*6+g.18400581C>T]<br>*6/[*6+g.18400193C>T] | *16/[g.18400841G>A]<br>*16/[g.18400349G>A]<br>*5/[*5+g.18400592C>T^]<br>*5/[*5+*44]<br>*5/[*5+*45]<br>*19/[*5+*34]<br>*5/[*5+g.18400841G>A]<br>*5/[*5+g.18400251G>C]<br>*5/[g.18400841G>A]<br>*6/[*4+g.18400124A>T]<br>*6/[*6+g.18400367G>A]<br>*7/[*4+g.18400581C>T] | *16/g.18400841G>A<br>*5/g.18400841G>A<br>*5/[*5 + g.18400592C>T^]<br>*5/[*5 + *34]<br>*5/[*5 + *40]<br>*7/[*4 + *41]<br>*4/[*4 + g.18400841G>A]<br>*5/[*16 + g.18400841G>A]<br>*6/[*6 + g.18400367G>C]<br>*4/[*6 + g.18400367G>T]<br>*5/g.18400206G>C<br>*1/g.18400841G>A |

Table S5. PharmCAT-required *NAT2* variants, the star allele they define when found alone, and in how many of the PharmCAT v3.0.1's 48 *NAT2* star allele definitions they are included.

| Variant | Defining Star Allele | Number of Star Alleles Containing Variant |
|---|---|---|
| rs1208 A | *4 | 31 |
| rs1801280 C | *5 | 8 |
| rs1799930 A | *34 | 17 |
| rs1799931 A | *40 | 3 |
| rs1801279 | *46 | 5 |

Table S6. Comparison of selected *NAT2* allele frequencies from the All of Us, PennMedicine BioBank, and UK Biobank 500k datasets (analyzed with PharmCAT v3.0.1) and data from the PharmVar *NAT2* Gene Focus inferred single nucleotide variant frequencies. (Papanikolaou *et al*. 2025, PMID: 41432551).

Abb.: AFR – African participants; AMR – admixed American participants; EUR – European participants; EAS – East Asian participants; SAS – South Asian participants; PMBB – PennMedicine BioBank; UKBB – UK Biobank

| Allele | Papanikolaou *et al*. | | Biobanks | | | |
|---|---|---|---|---|---|---|
| | Population/Geographical Description | Frequency in % | Population | All of Us Frequency in % | PMBB Frequency in % | UKBB 500k Frequency in % |
| *4 | West African | ~20 | AFR | 22 | 22 | 22 |
| | Brazilians<br>Native American | ~20<br>≥50 | AMR | 31 | 30 | 30 |
| | European | ~20 | EUR | 23 | 23 | 23 |
| | East Asia | ≥50 | EAS | 52 | 52 | 51 |
| | Central Asia | ≥50 | SAS | 21 | 19 | 21 |
| *5 | East and North Africa | 30-50 | AFR | 29 | 30 | 28 |
| | European | 30-50 | EUR | 43 | 43 | 42 |
| *6 | All continents | ~20 | AFR | 22 | 22 | 21 |
| | | ~20 | AMR | 18 | 20 | 26 |
| | | ~20 | EUR | 29 | 30 | 30 |
| | | ~20 | EAS | 27 | 28 | 27 |
| | | ~20 | SAS | 36 | 37 | 35 |
| *7 | Americas | ~12 | AMR | 12 | 11 | 8 |
| | Asia | ~12 | EAS | 16 | 16 | 17 |
| | | ~12 | SAS | 7 | 8 | 7 |
| *1 | Central and West Africa | more prevalent | AFR | 8 | 7 | 8 |
| | than in most other populations | - | AMR | 3 | 1 | 2 |
| | | - | EUR | 0.4 | 0.3 | 0.5 |
| | | - | EAS | 0.5 | 0.4 | 0.7 |
| | | - | SAS | 1.5 | 0.6 | 1.2 |
| *14 | sub-Saharan Africa | notable frequency | AFR | 8 | 7 | 9 |
| | other world populations | rarely | AMR | 0.8 | 0.9 | 1.4 |
| | | rarely | EUR | 0.03 | 0.04 | 0.02 |
| | | rarely | EAS | 0.005 | 0 | 0.021 |
| | | rarely | SAS | 0.04 | 0 | 0.03 |

Table S7. Comparison of selected *NAT2* diplotype frequencies from the All of Us, PennMedicine BioBank, UK Biobank 500k unphased and UK Biobank 200k phased datasets (analyzed with PharmCAT v3.0.1) and data from Malinga *et al*. 2025 (PMID: 39829327, Data from Table S5). The UKBB 200k phased data were used to prioritize ambiguous diplotypes in unphased data. Therefore, the All of Us, PMBB, and UKBB 500k datasets have only diplotype frequencies for the prioritized diplotype.

Abb.: SSA – Sub-Saharan African participants; AFR – African participants; AMR – admixed American participants; EUR – European participants; EAS – East Asian participants; SAS – South Asian participants; PMBB – PennMedicine Biobank; UKBB – UK Biobank

| Diplotype | Malinga et al. - Frequency in % | | | | | UKBB 200k phased - Frequency in % | | | | |
|---|---|---|---|---|---|---|---|---|---|---|
| | SSA | AMR | EUR | EAS | SAS | AFR | AMR | EUR | EAS | SAS |
| *4/*5 | 1.3 | 0.9 | 2.6 | 0.6 | 1.8 | 13.1 | 17.2 | 18.8 | 4.3 | 14.4 |
| *1/*16 | 9.7 | 21.6 | 17.3 | 3.0 | 11.3 | 0.06 | 0 | 0.03 | 0 | 0 |
| *5/*6 | 0.2 | 1.2 | 3.0 | 0 | 1.4 | 11.8 | 14.2 | 25.0 | 2.3 | 23.2 |
| *16/*34 | 10.8 | 11.7 | 18.9 | 3.0 | 21.5 | 0 | 0 | 0.001 | 0 | 0 |
| *5/*7 | 0 | 0.3 | 0 | 0.2 | 0 | 1.7 | 5.8 | 2.0 | 1.3 | 4.5 |
| *16/*40 | 1.1 | 4.1 | 2.2 | 0.8 | 4.3 | 0 | 0 | 0 | 0 | 0 |
| *4/*49 | - | - | - | - | - | 0 | 0 | 0.002 | 0 | 0.021 |
| *16/*41 | 0.1 | 0 | 0 | 0 | 0 | 0 | 0 | 0 | 0 | 0 |

| Diplotype | All of Us - Frequency in % | | | | | PMBB - Frequency in % | | | | | UKBB 500k - Frequency in % | | | | |
|---|---|---|---|---|---|---|---|---|---|---|---|---|---|---|---|
| | AFR | AMR | EUR | EAS | SAS | AFR | AMR | EUR | EAS | SAS | AFR | AMR | EUR | EAS | SAS |
| *4/*5<br>*1/*16 | 12.7 | 20.5 | 19.4 | 4.2 | 13.1 | 13.6 | 21.6 | 19.6 | 3.3 | 12.7 | 12.8 | 17.5 | 18.9 | 4.2 | 14.3 |
| *5/*6<br>*16/*34 | 12.5 | 12.6 | 24.9 | 2.2 | 24.2 | 13.3 | 13.1 | 25.5 | 2.9 | 24.4 | 11.6 | 14.5 | 24.8 | 2.5 | 24.1 |
| *5/*7<br>*16/*40 | 1.9 | 7.6 | 2.1 | 1.3 | 4.3 | 2.1 | 7.5 | 2.1 | 1.7 | 6.6 | 1.5 | 5.8 | 2.0 | 1.4 | 4.7 |
| *4/*49<br>*16/*41 | 0.01 | 0.011 | 0.001 | 0 | 0.148 | 0 | 0 | 0.002 | 0 | 0 | 0 | 0 | 0.002 | 0.042 | 0.038 |
